# Using Best-Worst Scaling to investigate younger adult Canadians’ preferences for COVID-19 vaccination and public health measures: an observational study

**DOI:** 10.1101/2022.02.18.22271180

**Authors:** Ovidiu Tatar, Ben Haward, Patricia Zhu, Gabrielle Griffin-Mathieu, Samara Perez, Gregory Zimet, Zeev Rosberger

**Affiliations:** Lady Davis Institute for Medical Research (LDI), Jewish General Hospital, Montreal, QC, Canada; Research Center, Centre Hospitalier de l’Université de Montréal (CRCHUM), Montreal, QC, Canada; Department of Psychiatry, McGill University, Montreal, QC, Canada; McGill University Health Center (MUHC), Montreal, QC, Canada; Indiana University School of Medicine, IN, USA; Departments of Psychology, Psychiatry and Oncology, McGill University, Montreal, QC, Canada

**Keywords:** COVID-19, Vaccination, Public health measures, Preferences, Best-Worst Scaling, Vaccine Acceptability, Younger adults

## Abstract

Containing the COVID-19 pandemic is dependent on compliance with public health recommendations and mandates which is lower in younger compared to older adults. Furthermore, younger adults have demonstrated lower uptake of COVID-19 vaccines. The aim of this study was to assess preferences for COVID-19 related preventive health measures and vaccination and to explore their association with COVID-19 vaccine acceptability. Canadians aged 18-39 years were invited to participate in a web-based survey in August 2021. We used the Best-Worst-Scale (BWS) methodology to collect and analyze preference data and multivariable binary logistic regression to estimate associations with vaccine acceptability. Based on 266 complete responses, we found strong preferences for physical distancing and wearing face masks, as compared to general hygiene and respiratory etiquette. High vaccine accessibility independent of the location, receiving successive doses of the same vaccine brand and higher vaccine uptake of people in younger adults’ social circle were highly preferred. Higher preferences for mandates requiring proof of vaccination and altruistic motives for vaccination were associated with vaccine acceptability. As the COVID-19 pandemic waxes and wanes, studies using larger, nationally representative samples are needed to replicate and validate these results to assess preferences for health behaviors corresponding to the latest recommendations. The use of this methodology could provide public health authorities with a unique opportunity to develop targeted, preference-based messaging that aligns with the latest guidelines to effectively encourage compliance and COVID-19 vaccine uptake.

## 1. Introduction

The COVID-19 pandemic is the greatest public health challenge of recent times. Canada alone has reported over 1.69 million cases and 28,500 deaths at the time of writing, and mitigation strategies have had far-reaching effects on our social and economic landscape (1). Non-pharmaceutical measures, such as mask wearing, physical distancing, and restrictions on schools and businesses, remain essential tools to contain the COVID-19 pandemic and reduce the strain on healthcare systems. The introduction of vaccines has further bolstered our ability to combat the pandemic and has likely saved countless lives. However, vaccine refusal and non-compliance to recommended public health measures pose significant barriers to containing and ultimately halting the pandemic, particularly in light of recent, more highly infectious variants such as Delta (2, 3).

Younger adults have been identified as a specific and concerning population in the ongoing effort to control the pandemic. This age group has a greater number of social contacts and is more likely to experience mild or asymptomatic infection (4). Subsequently, they are less likely to be aware of infection and to isolate from others (5). Furthermore, numerous population-based studies have identified an association between younger age and public health measure non-compliance (6, 7). Despite COVID-19 vaccines now being widely available, in Canada, younger adults, aged 18-39, have a lower rate of full vaccination (about 80%) than older adults, aged 60+ (90-95%) in Canada (8). This reflects the findings of multiple studies suggesting younger age is associated with greater COVID-19 vaccine hesitancy (9, 10). Considering that younger adults’ continue to be a vector for COVID-19 transmission, and even increasingly face severe consequences from infection with the rise of new variants (11), this is of great concern to public health authorities (12).

Attitudes and beliefs towards COVID-19 public health measures and vaccination vary across age groups, suggesting that the acceptability of, and compliance with, measures and recommendations will also vary with age (10, 13). A targeted approach is needed to clarify measures that will produce sufficient acceptance, compliance, and vaccine uptake in the 18-39 age group as the pandemic continues. Preferences are important indicators of behavior (14, 15), but only a handful of studies have used individual preferences for COVID-19 measures and vaccination to understand the public response to mitigation measures (14, 16-18), and none of these studies assessed preferences in younger Canadian adults. In the last decade, the Best-Worst Scaling (BWS) methodology (grounded in the microeconomic theory) has gained traction in evaluating preferences for interventions in healthcare (19, 20) but it has yet to be used in the context of the COVID-19 pandemic. Compared to using conventional multiple choice questions in which preferred attributes are selected only once, the BWS offers a more in-depth, nuanced understanding of preferences as it is based on the concept of utility trade-off (21, 22). Correspondingly, in BWS, participants are asked to identify the best (most preferred) and the worst (least preferred) item in multiple questions that contain a random list of usually three or four attributes (e.g., wearing face masks) and their corresponding three or four attribute-levels (e.g., when using public transportation or shopping’, at work or at school).

Using more advanced evaluation methods of public preferences could assist public health authorities in modifying and aligning guidelines, and in turn build trust, ensure acceptability, and increase compliance with mitigating strategies to address the COVID-19 pandemic. The objectives of this study were to advance our understanding of the preferences of Canadian younger adults (aged 18-39) for COVID-19 public health measures and vaccination; and to explore the associations between these preferences and COVID-19 vaccine acceptability.

## 2. Methods

### 2.1. Study design

In August 2021, we used a cross-sectional design and a web-based survey to collect data from Canadian adults. We provided the study details according to the Strengthening the Reporting of Observational Studies in Epidemiology (STROBE) recommendations (23). Ethical approval was obtained from the Research Ethics Board of the CIUSSS West-Central Montreal (Project ID 2022-2877).

### 2.2. Setting and participants

Canadian residents aged 18-39 years independent of their COVID-19 vaccination status were invited to complete the questionnaire in English. At the time of data collection COVID-19 vaccines were available for Canadians 12 years and older in all jurisdictions and the national uptake rate in the population of interest was about 70% (8). Data collection was facilitated by Dynata, an international survey company that used different platforms (e.g., direct email, smartphone app notifications, their own website) to invite participants. We used quotas for sex and province of residence (according to national census data) to ensure a balanced sample. After providing electronic consent, participants completed the questionnaire on smartphones or computers and were compensated according to Dynata’s rewards and points system (e.g., Amazon, Starbucks).

### 2.3. Variables

Sociodemographics included continuous (i.e., age) and categorical variables [gender province or territory; ethnicity; self-perceived visible minority (yes/no); influence of religion on health decisions (yes/no); language spoken at home (English, French, Other); post-secondary education attainment (yes/no); and income (ten thousand dollars increments)]. Variables with small cell count for some categories were re-categorized. Thus, province or territory was re-categorized into Western, Central, or Atlantic Canada. The nine categories used by Statistics Canada to measure self-reported ethnic origins (24) were re-categorized into North American, European, Asian, or Other (i.e., Caribbean, Latin, Central and South American, African). We used multiple validated categories (25) to measure gender identity that captures men and women’s socially constructed roles, identities and behaviors and retained for analyses three categories: male, female and gender diverse (i.e., Transgender Male/Trans Man/ Female-to-Male; Transgender Female/Trans Woman/ Male-to-Female; Genderqueer, neither exclusively male nor female; Other (open ended); and Choose not to disclose). For self-reported yearly family income in the year preceding the pandemic, we created three categories using 40- and 80-thousand-dollar cut-offs.

Informed by the literature search conducted by our team, we included measures that were associated with COVID-19 vaccine acceptability. We used the following dichotomous (yes/no) variables: healthcare professional status; caregiver of an elderly person; receipt of the flu vaccine in the last 12 months; history of testing positive for COVID-19 for oneself and friends/family. The validated 6-point-item (excellent to very poor) measure of self-perceived health status (26) was dichotomized into “excellent or very good” and “good or less”. Smoking history was captured by three categories: never; former; and current smoker.

We used the Precaution Adoption Process Model (PAPM) (27) to measure COVID-19 vaccine acceptability and participants selected one of the five nominal intention stages: *unengaged*; *undecided*; *decided not*; *decided to vaccinate*; and *vaccinated*. The outcome was dichotomized into vaccine acceptors (stages *decided to* and *vaccinated*) and vaccine hesitant/rejectors (*unengaged, undecided* and *decided not*).

Preferences for preventive public health measures and COVID-19 vaccination were measured using the case 2 BWS methodology (19, 20). Two domains of preferences were determined for preventive public health measures (A – preventive health behaviors; B – government mandates) and two for vaccination (C – immunization specific; D – motives for vaccination). For each domain, we defined attributes that reflect high-order preferences within a domain and within each attribute we used attribute-levels to measure preferences for an attribute. We selected attributes and attribute-levels for each domain based on public health measures and COVID-19 vaccination recommendations available on the website of the Government of Canada and government mandates (e.g., curfew, travelling limitations) (28) at the time of study conception (April-June 2021). Additionally, extant preferences (e.g., altruistic vaccination motives, self-interested motives) were chosen based on relevant literature (29, 30) and concepts included in media campaigns designed to improve vaccine uptake. To measure preferences, a total of 57 questions were answered, 16 questions in each of the domains A, B and D and 9 questions in domain C (Table 1, sample questions).

**Table 1.** Sample questions for domain A (preventive health behaviours) and D (vaccination motives)

| Question 1 (out of 16) |  |  |
| --- | --- | --- |
| LEAST preferred | Options | MOST preferred |
|  | ...frequently washing my hands for at least 20 seconds with soap and water or using hand sanitizers |  |
|  | ...avoiding exposure to closed or crowded spaces |  |
|  | ...wearing a face mask when using public transportation or shopping |  |

I would get the COVID-19 vaccine to...
| Question 1 (out of 16) |  |  |
| --- | --- | --- |
| LEAST preferred | Options | MOST preferred |
|  | ...protect my family |  |
|  | ...allow others to go back to school |  |
|  | ...allow myself to socialize at restaurants, bars, etc. |  |

We created a separate set of questions for each domain in two phases. First, we defined the attributes and their corresponding attribute-levels, and second, we used the orthogonal main effect design methodology recommended by Aizaki, H and Fogarty, J (2019) (31) and the R software packages “DoE.base” (32) and “support.BWS2” (33) to generate the full set of questions for each of the four domains. For each domain, participants answered a distinct set of questions (generated in R using the functions “oa.design” and “bws2.questionnaire”) that account for the number of attributes and their corresponding attribute-levels. The domains A, B, and D measured three attributes with four attribute-levels each and domain C measured four attributes with three attribute-levels each. Each question within a domain consisted of an equal number of randomly allocated attribute-levels and participants selected the best or the worst preferred attribute-level. To mitigate the possibility of response bias, the order of questions within a domain and the order of domains within the questionnaire were randomized for each participant.

### 2.4. Statistical analysis

To analyze BWS preference data, we used the counting and the modelling approaches described by Aizaki et al. (2019) to assess preferences (31). In line with the counting approach, within each domain we calculated the best-minus-worst (BW) total score for each attribute and attribute level. For each observation, the BW score (BWs) is calculated by subtracting the number of times an attribute-level (or attribute) X is selected as the worst from the number of times an attribute-level (or attribute) X is selected as the best among all the questions in a domain. The total BWs is obtained by summing the scores for all observations and higher scores reflect higher preference. We calculated scores using the function “bws2.count” from the package “support.BWS2” in R.

For the modelling approach, we used conditional logistic regression to model preferences as a function of the sum of the utility of attributes and attribute-levels (31). We used the marginal model based on the assumption that respondents evaluated all attribute-levels both when choosing the best and the worst attribute-level in each question. In each of the four models (corresponding to domains A to D), one attribute and one attribute-level (per attribute) that had the lowest total BWs were omitted from the utility function and treated as reference categories. We estimated the odds ratios (OR) and 95% confidence intervals (CI) using the “clogit” function which is part of the “survival” package in R (34).

To explore the correlates of COVID-19 vaccine acceptability we used binary logistic regression and reported OR and their 95% CI. Sociodemographics and other health related behaviors that were significantly associated with vaccine acceptability in bivariate analyses were included in the multivariable model. All variables corresponding to attributes with a positive total BWs from all the four domains were simultaneously included in the model. We used the following logistic regression model diagnostic criteria: 1) Rank Discrimination Index C whereby higher C values indicated better model ability to classify individuals correctly into groups according to their outcome, 2) Cessie–van Houwelingen goodness-of-fit test whereby p > 0.05 suggests no evidence to reject a good fit (35), and 3) Variation Inflation Factor (VIF) with a cutoff value of <10 for ruling out collinearity issues (36).

The sample size was calculated based on the work of Peduzzi et al., (1995) who recommended a minimum of 10 observations per variable to adequately power binary logistic regression models (37). At the time of study conception, we estimated a 60% vaccine uptake and calculated that a sample of 250 participants would be required based on the formula N = 10k/p where N = minimum number of observations needed, k = number of predictor variables and p = smallest of the proportion in the binary model (i.e., N = 10*10/0.4). Based on the data provided by Cheung et al., (2016) in their systematic review, we calculated that studies using the case 2 BWS methodology in healthcare recruited an average of 316 participants with a median of 162 and range 16 to 1296 participants. Therefore, we estimated that a sample of about 250 would be adequate to conduct both BWS and multivariable logistic regression analyses. We conducted all analyses using RStudio and the R software v. 4.0.5.(38)

## 3. Results

Between August 6-18, 2021, 308 participants enrolled in the survey, 26 (8.4%) abandoned and 16 (6%) were terminated during completion of the survey by Dynata’s internal mechanisms to identify inattentive responders. The final dataset consisted of 266 observations with no missing data as questions could not be skipped. Vaccine hesitant participants (n=68; 25.6%) were in the PAPM decision stages *unengaged* (n=20; 7.5%); *undecided* (n=30; 11.3%) or *decided not* (n=18; 6.8%) while vaccine acceptors (n=198; 74.4%) included stages *decided to* (n=20; 7.5%) and *vaccinated* (n=178; 66.9%).

### 3.1. Preferences for preventive public health measures

At the attribute level, the most preferred preventive health behavior was physical distancing (BWs= 124) followed by wearing face masks (BWs = 32) and the least preferred was respecting general hygiene and respiratory etiquette (BWs = -156). Physical distancing (OR = 1.10) and wearing face masks (OR = 1.07) were preferred over respecting general hygiene and respiratory etiquette. As shown by the descending order of all attribute-level BWs in domain A (preventive health behaviours), the most preferred behavior was “avoiding exposure to closed or crowded spaces” (BWs = 88) and the least preferred was “wearing a face mask in open spaces such as the park or on the street” (BWs = - 133). With respect to physical distancing, the most preferred behavior was avoiding exposure to closed or crowded spaces (OR = 1.13). Participants preferred wearing face masks when using public transportation or shopping (OR= 1.15) and when two meters distancing cannot be kept (OR = 1.13) over wearing masks in open spaces. To prevent the spread of the virus, participants preferred adequate hand washing (OR = 1.10) compared to respecting the recommended sneezing etiquette. (See Table 2).

**Table 2.** Preferences for attributes and attribute levels corresponding to domain A (preventive health behaviours)

| **Attributes** | **BWs** | **OR** | **95% CI** |
| --- | --- | --- | --- |
| Physical distancing | 124 | **1.10** | **1.05; 1.16** |
| Wearing face masks | 32 | **1.07** | **1.01; 1.13** |
| General hygiene and respiratory etiquette | -156 | ref |  |
| **Levels for attribute: physical distancing** |  |  |  |
| ...avoiding exposure to closed or crowded spaces | 88 | **1.13** | **1.04; 1.22** |
| ...limiting contact with those at higher risk such as the elderly and those with a weaker immune system | 66 | 1.08 | 0.99; 1.17 |
| ...maintaining a physical distance of 2 meters from people outside of my household | 33 | 1.00 | 0.93; 1.09 |
| ...avoiding non-essential travel outside of Canada | -63 | ref |  |
| **Levels for attribute: wearing face masks** |  |  |  |
| ...wearing a face mask when using public transportation or shopping | 75 | **1.15** | **1.07; 1.25** |
| ...wearing a face mask in situations when I cannot keep a 2-meter distance from others | 65 | **1.13** | **1.04; 1.22** |
| ...wearing a face mask at work or at school | 25 | 1.04 | 0.96; 1.12 |
| ...wearing a face mask in open spaces such as at the park or on the street | -133 | ref |  |
| **Levels for attribute: general hygiene and respiratory etiquette** |  |  |  |
| ...frequently washing my hands for at least 20 seconds with soap and water or using hand sanitizers | 6 | **1.10** | **1.02; 1.19** |
| ...regularly disinfecting surfaces I frequently touch with my hands | -33 | 1.01 | 0.94; 1.10 |
| ...avoiding touching my eyes, nose, or mouth with unwashed hands | -59 | 0.96 | 0.89; 1.04 |
| ...coughing and sneezing into a tissue or the bend of my arm, not my hand | -70 | ref |  |

**Note:** in bold significant 95% confidence interval (CI) of odds ratios (OR); ref= the reference category for levels and attributes; BWs = best-worst score.

The most preferred government mandates attribute was the request to provide proof of health (BWs = 960) followed by imposing travelling limitations (BWs = 199) and the least preferred were measures to reduce the exposure to the virus (BWs=-1159). Preferences for the first two attributes were significantly higher than for the last one (OR = 2.18 and OR = 1.65 respectively). Among all attribute-levels studied in domain B (government mandates), the most preferred was the request to provide vaccination proof when entering Canada (BWs = 358) and the least preferred were evening or overnight stay at home orders (BWs = -415). While requiring proof of vaccination for entering Canada was more popular (OR = 1.31) than regular proof of a negative COVID test to attend work or school, the reverse was true for mandatory proof of vaccination to return to work or school (OR = 0.90). Participants preferred mandatory testing measures (OR = 1.41) and quarantine after arriving in Canada (OR = 1.10) and disliked restrictions on travel within provinces (OR = 0.81) compared to restriction on travel between provinces. Compared to curfew, preferences were higher for mandatory remote work or online classes (OR = 1.47) and lower for reduced hours for non-essential businesses (OR = 0.84). (See Table 3).

**Table 3.** Preferences for attributes and attribute levels corresponding to domain B (government mandates)

| **Attributes** | **BWs** | **OR** | **95% CI** |
| --- | --- | --- | --- |
| Requiring proof of health | 960 | **2.18** | **2.06; 2.30** |
| Traveling limitations | 199 | **1.65** | **1.56; 1.74** |
| Reducing exposure to the virus | -1159 | ref |  |
| **Levels for attribute: requiring proof of health** |  |  |  |
| ...proof of vaccination for entering Canada | 358 | **1.31** | **1.21; 1.42** |
| ...proof of vaccination to attend large gatherings such as sports, music, and religious events | 244 | 1.00 | 0.93; 1.09 |
| ... proof of vaccination to return to work or school | 195 | **0.90** | **0.83; 0.98** |
| ...regular proof of negative COVID tests to attend work or school | 163 | ref |  |
| **Levels for attribute: traveling limitations** |  |  |  |
| ... mandatory COVID test for any air travel | 206 | **1.41** | **1.30; 1.53** |
| ... mandatory quarantine after arriving in Canada | 93 | **1.10** | **1.01; 1.19** |
| ... restrictions on travel within provinces | -47 | **0.81** | **0.75; 0.88** |
| ... restrictions on travel between provinces | -53 | ref |  |
| **Levels for attribute: reducing exposure to the virus** |  |  |  |
| ... mandatory remote work for non-essential workers or online classes for students | -119 | **1.47** | **1.36; 1.59** |
| ... limitations on the number of people that can meet for socializing or leisure purposes | -258 | 1.08 | 1.00; 1.17 |
| ... reduced hours for non-essential businesses | -367 | **0.84** | **0.77; 0.91** |
| ... Evening or overnight stay at home orders (curfew) | -415 | ref |  |

**Note:** in bold significant 95% confidence interval (CI) of odds ratios (OR); ref= the reference category for levels and attributes; BWs = best-worst score.

### 3.2. Preferences for COVID-19 vaccination

The most preferred immunization specific attribute was vaccine accessibility (BWs = 231) followed by the vaccination status of other people (BWs=92) and vaccine dosing (BWs=9) and the least preferred was the pairing of flu vaccination (BWs=-332) with the COVID-19 vaccine. Higher preferences were expressed for the first three attributes compared to the last (OR=1.61; OR=1.43 and OR = 1.33 respectively). Among all attribute-levels in domain C (immunization specific), both the most (receiving 2 doses of the same vaccine, BWs=133) and the least preferred (receiving two doses of two different brands, BWs=-123) were captured by the vaccine dosing attribute. Possible drivers of vaccine acceptability were high vaccine uptake amongst close others (85% vs. 40%, OR=1.15) and the availability of the same vaccine brand for the second dose compared to different brands (OR=1.55) (Table 4).

**Table 4.** Preferences for attributes and attribute levels corresponding to domain C (immunization specific)

| **Attributes** | **BWs** | **OR** | **95% CI** |
| --- | --- | --- | --- |
| Vaccine accessibility | 231 | **1.61** | **1.49; 1.75** |
| Vaccination status of other people | 92 | **1.43** | **1.32; 1.55** |
| Vaccine dosing | 9 | **1.33** | **1.23; 1.45** |
| Comparison vaccines | -332 | ref |  |
| **Levels for attribute: vaccine accessibility** |  |  |  |
| ...I could get vaccinated at a doctor's office/clinic | 85 | 1.03 | 0.94; 1.13 |
| ...I could get vaccinated at a pharmacy | 83 | 1.02 | 0.93; 1.12 |
| ...I could get vaccinated at a vaccination site | 63 | ref |  |
| **Levels for attribute: vaccination status of other people** |  |  |  |
| ...85% of my family, friends and acquaintances were already vaccinated | 71 | **1.15** | **1.05; 1.26** |
| ...60% of my family, friends and acquaintances were already vaccinated | 31 | 1.00 | 0.91; 1.10 |
| ...40% of my family, friends and acquaintances were already vaccinated | -10 | ref |  |
| **Levels for attribute: vaccine dosing** |  |  |  |
| ...I could get vaccinated with 2 doses of the same vaccine | 133 | **1.55** | **1.42; 1.71** |
| ...I could get vaccinated with only one dose of a vaccine | -1 | 0.99 | 0.90; 1.08 |
| ...I could get vaccinated with 2 doses of two different vaccine brands | -123 | ref |  |
| **Levels for attribute: integration with flu vaccination** |  |  |  |
| ...I were to receive both the COVID-19 vaccine and the flu vaccine at the same time | -99 | 1.04 | 0.95; 1.14 |
| ...I were to receive the COVID-19 vaccine and the flu vaccine but NOT at the same time | -116 | 0.98 | 0.89; 1.08 |
| ... I were to only receive the COVID-19 vaccine and not the flu vaccine | -117 | ref |  |

**Note:** in bold significant 95% confidence interval (CI) of odds ratios (OR); ref= the reference category for levels and attributes; BWs = best-worst score.

We found that altruism motives (BWs=1641) were of higher importance than receiving the vaccine to reduce the impact of the pandemic on the society (BWs=-535) or self-interested motives (BWs=-1106). Both altruism motives (OR=2.79) and impact on the society (OR=1.24) were preferred over self-interested motives. Among all attribute-level motives in domain D (motives for vaccination), protecting one’s family (BWs=490) was the most and going to the gym (BWs=-395) the least important motive. Among altruism motives, protecting one’s family was more important (OR=1.22) than protecting friends, classmates or coworkers while protecting the whole community was less important (OR=0.90). At a societal level, reducing the burden on the healthcare system was preferred (OR=1.50) over facilitating large social gatherings. Among self-interested motives, travelling without restrictions (OR=1.14) and socializing at bars, restaurants (OR=1.11) were of higher importance than going to the gym. (See Table 5).

**Table 5.** Preferences for attributes and attribute levels corresponding to domain D Motives for vaccination)

| **Attributes** | **BWs** | **OR** | **95% CI** |
| --- | --- | --- | --- |
| Altruism motives | 1641 | **2.79** | **2.63; 2.95** |
| Impact on society | -535 | **1.24** | **1.17; 1.31** |
| Self-interested motives (leisure) | -1106 | ref |  |
| **Levels for attribute: altruism motives** |  |  |  |
| ...protect my family | 490 | **1.22** | **1.12; 1.33** |
| ...protect vulnerable persons such as children, the elderly, and the chronically ill | 419 | 1.02 | 0.94; 1.11 |
| ...protect everyone in my community | 367 | **0.90** | **0.83; 0.98** |
| ...protect my friends, classmates, or coworkers | 365 | ref |  |
| **Levels for attribute: impact on society** |  |  |  |
| ...help reduce the burden on the healthcare system | 47 | **1.50** | **1.38; 1.62** |
| ... allow others to go back to school | -152 | 0.96 | 0.89; 1.04 |
| ...allow others to go back to work | -170 | 0.92 | 0.85; 1.00 |
| ...allow others to attend large gatherings such as sports, music, and religious events | -260 | ref |  |
| **Levels for attribute: self-interested motives (leisure)** |  |  |  |
| ...allow myself to travel without restrictions | -221 | **1.14** | **1.05; 1.24** |
| ... allow myself to socialize at restaurants, bars, etc. | -232 | **1.11** | **1.02; 1.20** |
| ...allow myself to attend large gatherings such as sports, music, and religious events | -258 | 1.05 | 0.96; 1.14 |
| ...allow myself to go to the gym | -395 | ref |  |

**Note:** in bold significant 95% confidence interval (CI) of odds ratios (OR); ref= the reference category for levels and attributes; BWs = best-worst score.

### 3.3. Correlates of vaccine acceptability

In bivariate analyses, age (OR=1.08) and Asian ethnicity (OR=3.50) were associated with higher odds of vaccine acceptability. Influence of religion on health decisions (OR=0.47), being a healthcare professional (OR=0.34) or a caregiver of an elderly person (OR=0.43) or having tested positive for COVID (OR= 0.43) were associated with lower vaccine acceptability (Table 6). In multivariable analysis, the association of age (AOR=1.12) and Asian ethnicity (AOR=8.37) remained unchanged. Higher preferences for mandates related to providing proof of health (e.g., vaccination) or altruism motives (e.g., protecting one’s family) were associated with higher odds of vaccine acceptability (AOR=1.16 and AOR=1.06 respectively) (Table 7).

**Table 6.**
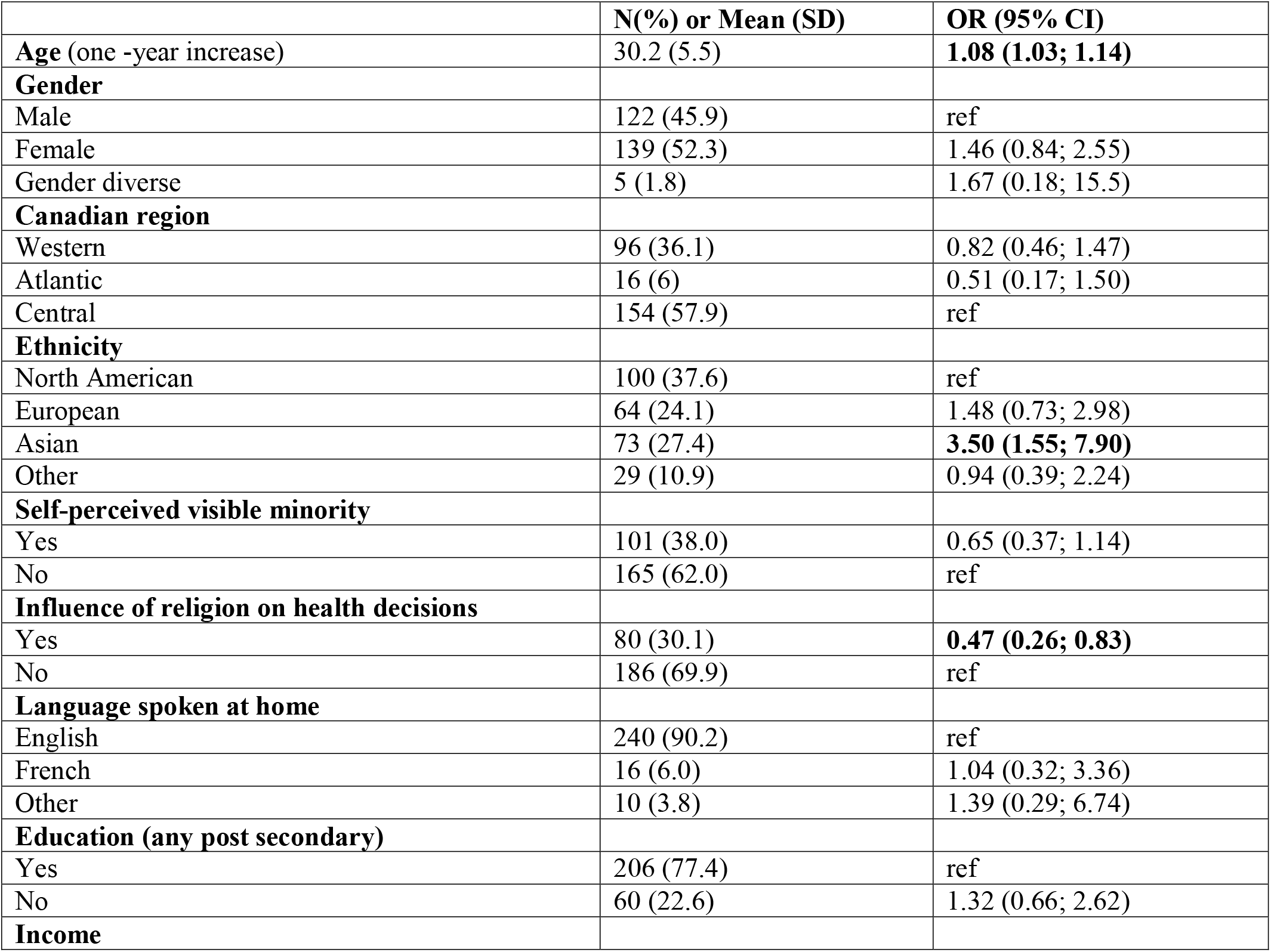
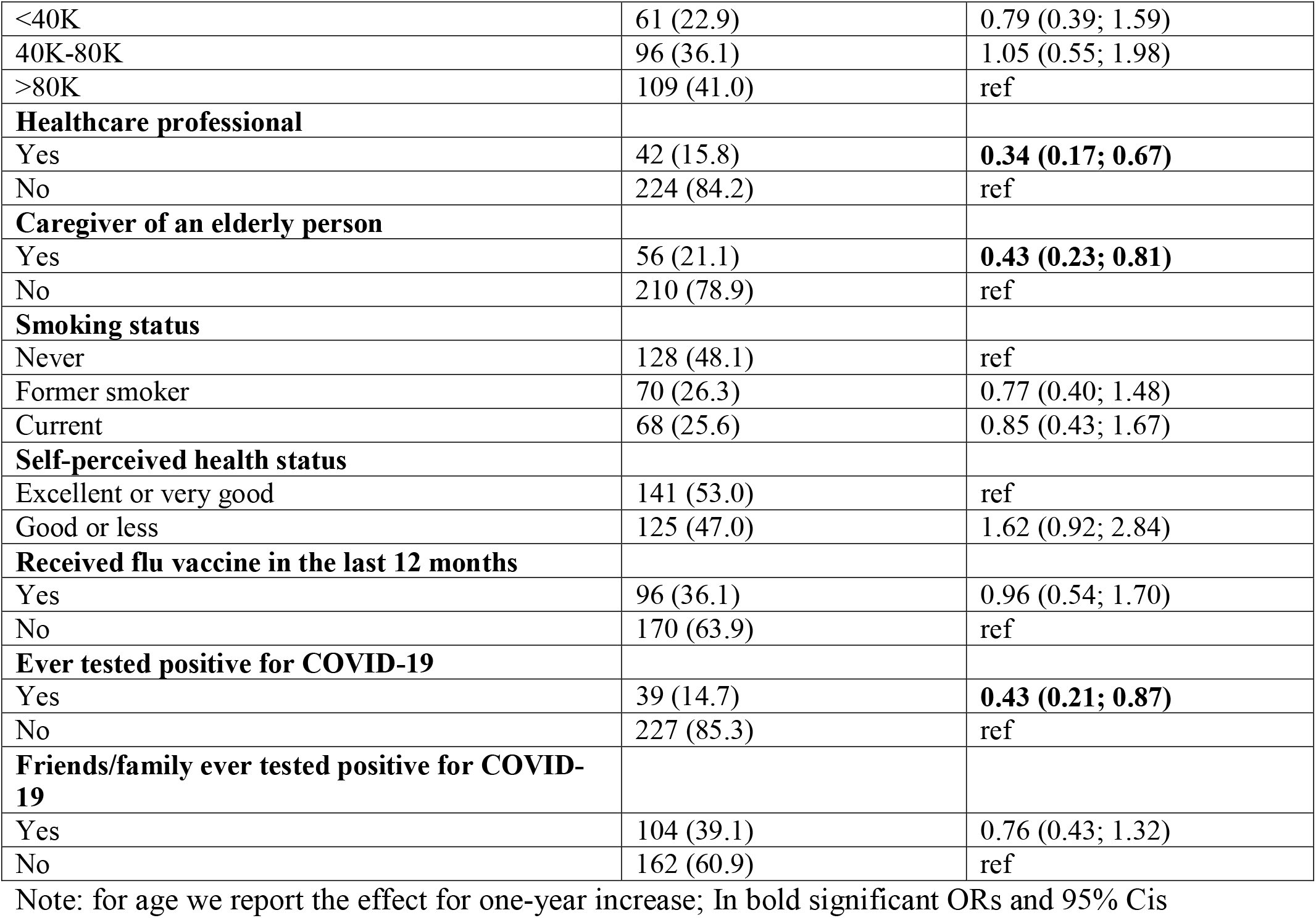
Sociodemographics and health behaviors. Bivariate associations with vaccine acceptability (n=266)

**Table 7.** Results of multivariable logistic regression (n=266)

|  | **AOR (95% CI)** |
| --- | --- |
| **Age** | **1.12 (1.05; 1.21)** |
| **Ethnicity** |  |
| North American | ref |
| European | 1.50 (0.62; 3.67) |
| Asian | **8.37 (2.88; 24.32)** |
| Caribbean, Latin, Central and South American, African | 2.13 (0.71; 6.38) |
| **Influence of religion on health decisions** |  |
| Yes | 0.80 (0.35; 1.79) |
| No | ref |
| **Healthcare professional** |  |
| Yes | 0.55 (0.19; 1.58) |
| No | ref |
| **Caregiver of an elderly person** |  |
| Yes | 0.79 (0.30; 2.09) |
| No | ref |
| **Ever tested positive for COVID-19** |  |
| Yes | 1.38 (0.45; 4.27) |
| No | ref |
| **BWS Attributes** |  |
| Physical distancing | 0.94 (0.87; 1.02) |
| Wearing face masks | 0.95 (0.89; 1.01) |
| Vaccination status of other people | 1.07 (0.94; 1.22) |
| Vaccine accessibility | 1.13 (0.97; 1.31) |
| Vaccine dosing | 0.94 (0.80; 1.11) |
| Requiring proof of health | **1.16 (1.09; 1.25)** |
| Traveling limitations | 1.00 (0.92; 1.09) |
| Altruism | **1.06 (1.02; 1.11)** |

Note: for age we report the effect for one-year increase; for BWS attributes we report the effect for one unit increase in the BW score. In bold significant ORs and 95% CIs Rank Discrimination Index C = 0.85; Cessie–van Houwelingen goodness-of-fit test p=0.21; VIF range: 1.19 to 2.74.

## 4. Discussion

Considering that adequate compliance to preventive health behaviour recommendations in younger adults is critical to contain the pandemic, we aimed to investigate their preferences for public health measures and vaccination and to explore their association with COVID-19 vaccine acceptability. This was captured using Best-Worst Scaling methodology, which has not previously been used in the context of the COVID-19 pandemic.

Physical distancing, and in particular “avoiding exposure to closed or crowded spaces”, was preferred, suggesting that for most younger adults, restrictions on larger gatherings are favorable policies. This is surprising, as other studies investigating younger adults’ attitudes and beliefs towards, and compliance with physical distancing show this population are less compliant with such measures (6, 7, 13). Unlike these studies, our methodology required participants to consider the utility trade-offs of multiple attributes when evaluating a measure. Our results could posit that, compared to evaluating attribute levels using conventional, multiple-choice questions, younger adults may be more likely to endorse physical distancing when repeatedly assessing its value amongst attribute levels (utility trade-offs). Considering that adequate respiratory etiquette and hand hygiene are effective in reducing the incidence of respiratory tract infections (39) these behaviours have been strongly encouraged by public health authorities since the beginning of the pandemic. It is thereby concerning that we observed relative lower preference for these behaviours in our sample, although consistent with research showing that younger age is associated with reduced handwashing (40).

With regard to government mandates, younger adults showed a strong preference for requiring proof of health through vaccination or negative COVID-19 test. Importantly, multivariable analyses also demonstrated that this preference was associated with a greater likelihood of vaccine acceptance. However, our results are also reminiscent of the barriers to implementing these policies. For example, in bivariate analyses, it is somewhat surprising that being a healthcare professional or caretaker for the elderly was a correlate of vaccine hesitancy, although consistent with the results published by Head et al. (41). This may be related to issues faced in Canada and the United States, where mandated vaccination for healthcare professionals has caused considerable controversy. We also found that testing positive was a correlate of vaccine hesitancy. This may be linked to the idea that those who had the virus, (and perhaps themselves more riskier individuals) have greater immunity than from the vaccine, which has been reported widely in the media (42). It is unclear if these results are worthy of attention as they did not hold in the multivariable analyses, and confirmation in future studies is needed. Despite this, our results suggest that for the most part, younger adult Canadians are willing to provide proof of health through vaccination or testing, and that backlash to these policies will most likely emanate from those who are unvaccinated. Additionally, aversion to mandatory limitations to gathering, in-person school and work, and non-essential businesses, might further suggest that most younger adults prefer mandates that require vaccination or other proof of health if they can continue their usual activities.

We found no significant difference in preference for vaccination at a doctor’s office, pharmacy, or vaccination site. This might suggest that tailored vaccine administration models for younger adults should focus less on which sites are optimal, and rather prioritize flexibility by having the vaccine available in many different settings. Participants preferred being vaccinated with two doses of the same vaccine over receiving only a single dose or two doses of different vaccine brands. As evidence emerges that “mix-and-match” booster shot strategies could be effective in providing sustained protection against COVID-19 (43), acceptance of this strategy for both initial vaccinations, and potentially also booster shots, might be lower in younger adults. Policy makers should factor this into implementation of such strategies and targeted communications should address concerns about vaccine mixing. While the joint administration of COVID-19 and influenza vaccination may be a promising solution to increase coverage for both vaccines (44), our results suggest it would not motivate younger adults to take a COVID-19 vaccine and, like vaccine mixing, may require interventions to improve acceptability. Similar to the results of Leng et al. (45) who found that increased vaccine uptake was associated with acceptability in unvaccinated individuals, we found that younger adults were more likely to prefer receiving a COVID-19 vaccine if 85% of their family, friends, and acquaintances had already received it. This finding could reflect “free riding” in which an individual waits to view the consequences of others’ behaviour before acting, while benefitting from the externalities, such as herd-immunity, produced by those actions (46). This can be hugely detrimental to vaccine uptake, as widespread free riding behaviour decreases the number of individuals proactively seeking vaccination, thus reinforcing the hesitancy of free riders.

Corresponding to other studies showing that altruistic reasons for vaccination are an effective way to promote vaccine acceptance (47, 48), we found a strong association between altruistic motives, and vaccine acceptance. Our results indicate that using altruistic motives to promote vaccination in this population might be effective, particularly when targeted towards the protection of one’s family. Self-interested motives for COVID-19 vaccination, despite being highly prevalent in current public health messaging, were not preferred by our sample, suggesting that messages promoting external benefits of vaccination (e.g., altruistic motives, reducing the burden on the healthcare system) may be more suitable for this age group.

### 4.1 Study Strengths and Limitations

Being limited by our sample size, we did not conduct sub-group analyses to assess preferences for preventive health behaviours in groups that are extremely important from the public health perspective such as healthcare professionals. Studies using BWS in larger, representative samples are needed to confirm our results and to examine differences in these subgroups. While we tried to anticipate emerging public health measures and recommendations, the COVID-19 pandemic is a rapidly evolving situation, and new challenges have emerged since the inception of our study. Future studies using this methodology would benefit from surveying the updated situation and considering preferences for innovative, emerging solutions such as mobile approaches to vaccine administration (49). Further, including the BWS methodology in web-based surveys is facilitated by special R packages and enables an advanced understanding of preferences based on the utility trade-off principle. In our opinion, using multiple categories to measure gender is insufficient to capture the influence of gender identity on health behaviors in studies that do not benefit from large samples. A possible mitigation strategy is to include ordinal scales to measure femininity and masculinity as recommended by Magliozzi et al.(50).

## 5. Conclusions

Using the Best-Worst-Scaling methodology in a younger adult population (aged 18-39), our findings provide a fine-tuned insight into the preferences of this age group whose adherence to public health recommendations and uptake of vaccines is critical to contain the pandemic. Our findings could inform public health authorities in aligning evidence-based guidelines with public preferences, to promote compliance with preventive measures and increase COVID-19 vaccination rates, including for possible upcoming booster shots.

## Supporting information

STROBE Reporting Checklist

## Data Availability

All data produced in the present study are available upon reasonable request to the authors

## REFERENCES

1. Government of Canada. Coronavirus disease 2019 (COVID-19): Epidemiology update 2021 [Available from:https://health-infobase.canada.ca/covid-19/epidemiological-summary-covid-19-cases.html#a2].

2. Dror AA, Eisenbach N, Taiber S, et al. Vaccine hesitancy: the next challenge in the fight against COVID-19. European Journal of Epidemiology,. 2020;35(8):775–9.doi: 10.1007/s10654-020-00671-y

3. Kupferschmidt K, Wadman M. Delta variant triggers new phase in the pandemic. Science. 2021;372(6549):1375–6.doi: 10.1126/science.372.6549.1375

4. Sah P, Fitzpatrick MC, Zimmer CF, et al. Asymptomatic SARS-CoV-2 infection: A systematic review and meta-analysis. Proc Natl Acad Sci U S A. 2021;118(34).doi: 10.1073/pnas.2109229118

5. Boehmer TK, DeVies J, Caruso E, et al. Changing Age Distribution of the COVID-19 Pandemic - United States, May-August 2020. MMWR Morb Mortal Wkly Rep. 2020;69(39):1404–9.doi: 10.15585/mmwr.mm6939e1

6. Valenti GD, Faraci P. Identifying Predictive Factors in Compliance with the COVID-19 Containment Measures: A Mediation Analysis. Psychology Research and Behavior Management. 2021;Volume 14:1325–38.doi: 10.2147/prbm.S323617

7. Coroiu A, Moran C, Campbell T, et al. Barriers and facilitators of adherence to social distancing recommendations during COVID-19 among a large international sample of adults. PLoS One. 2020;15(10):e0239795.doi: 10.1371/journal.pone.0239795

8. Government of Canada. COVID-19 vaccination in Canada 2021 [Available from:https://health-infobase.canada.ca/covid-19/vaccination-coverage/].

9. Afifi TO, Salmon S, Taillieu T, et al. Older adolescents and young adults willingness to receive the COVID-19 vaccine: Implications for informing public health strategies. Vaccine. 2021;39(26):3473–9.doi: 10.1016/j.vaccine.2021.05.026

10. Ogilvie GS, Gordon S, Smith LW, et al. Intention to receive a COVID-19 vaccine: results from a population-based survey in Canada. BMC Public Health. 2021;21(1):1017.doi: 10.1186/s12889-021-11098-9

11. Centers for Disease Control and Prevention. COVID-19 Associated Hospitalizations by Age 2021 [Available from:https://gis.cdc.gov/grasp/COVIDNet/COVID19_5.html].

12. Government of Canada. Update on COVID-19 in Canada: Epidemiology and modelling, September 3rd, 2021. 2021.

13. Lang R, Benham JL, Atabati O, et al. Attitudes, behaviours and barriers to public health measures for COVID-19: a survey to inform public health messaging. BMC Public Health. 2021;21(1):765.doi: 10.1186/s12889-021-10790-0

14. Eshun-Wilson I, Mody A, McKay V, et al. Public Preferences for Social Distancing Policy Measures to Mitigate the Spread of COVID-19 in Missouri. JAMA Netw Open. 2021;4(7):e2116113.doi: 10.1001/jamanetworkopen.2021.16113

15. Borriello A, Master D, Pellegrini A, et al. Preferences for a COVID-19 vaccine in Australia. Vaccine. 2021;39(3):473–9.doi: 10.1016/j.vaccine.2020.12.032

16. Manipis K, Street D, Cronin P, et al. Exploring the Trade-Off Between Economic and Health Outcomes During a Pandemic: A Discrete Choice Experiment of Lockdown Policies in Australia. Patient. 2021;14(3):359–71.doi: 10.1007/s40271-021-00503-5

17. Chorus C, Sandorf ED, Mouter N. Diabolical dilemmas of COVID-19: An empirical study into Dutch society’s trade-offs between health impacts and other effects of the lockdown. PLoS One. 2020;15(9):e0238683.doi: 10.1371/journal.pone.0238683

18. Reed S, Gonzalez JM, Johnson FR. Willingness to Accept Trade-Offs Among COVID-19 Cases, Social-Distancing Restrictions, and Economic Impact: A Nationwide US Study. Value Health. 2020;23(11):1438–43.doi: 10.1016/j.jval.2020.07.003

19. Cheung KL, Wijnen BF, Hollin IL, et al. Using Best-Worst Scaling to Investigate Preferences in Health Care. Pharmacoeconomics. 2016;34(12):1195–209.doi: 10.1007/s40273-016-0429-5

20. Muhlbacher AC, Kaczynski A, Zweifel P, et al. Experimental measurement of preferences in health and healthcare using best-worst scaling: an overview. Health Econ Rev. 2016;6(1):2.doi: 10.1186/s13561-015-0079-x

21. Finn A, Louviere JJ. Determining the Appropriate Response to Evidence of Public Concern: The Case of Food Safety. Journal of Public Policy & Marketing. 1992;11(2):12–25.doi: 10.1177/074391569201100202

22. Szeinbach SL, Barnes JH, McGhan WF, et al. Using Conjoint Analysis to Evaluate Health State Preferences. Drug Information Journal. 1999;33(3):849–58.doi: 10.1177/009286159903300326

23. Vandenbroucke JP, von Elm E, Altman DG, et al. Strengthening the Reporting of Observational Studies in Epidemiology (STROBE): explanation and elaboration. PLoS Med. 2007;4(10):e297.doi: 10.1371/journal.pmed.0040297

24. Canada S. List of ethnic origins 2016 [Internet]. 2016 [updated October 25, 2017. Available from:https://www23.statcan.gc.ca/imdb/p3VD.pl?Function=getVD&TVD=402936].

25. National LGBT Health Education Center. Collecting Sexual Orientation and Gender Identity Data in Electronic Health Records [Internet]. 2016 [cited 2021 November 2]. Available from:https://tinyurl.com/y66ln8do].

26. Bowling A. Just one question: If one question works, why ask several? J Epidemiol Community Health. 2005;59(5):342–5.doi: 10.1136/jech.2004.021204

27. Weinstein S, Sandman P, Blalock S. The Precaution Adoption Process Model. 4 ed. San Francisco, CA: Jossey-Bass; 2008. 123–48 p.

28. Government of Canada. Coronavirus disease (COVID-19): Prevention and risks [Internet]. 2021 [updated October 28, 2021; cited 2021 November 2]. Available from:https://www.canada.ca/en/public-health/services/diseases/2019-novel-coronavirus-infection/prevention-risks.html?topic=tilelink].

29. Hershey JC, Asch DA, Thumasathit T, et al. The Roles of Altruism, Free Riding, and Bandwagoning in Vaccination Decisions. Organizational Behavior and Human Decision Processes. 1994;59(2):177–87.doi: 10.1006/obhd.1994.1055

30. Cucciniello M, Pin P, Imre B, et al. Altruism and vaccination intentions: Evidence from behavioral experiments. Soc Sci Med. 2021:114195.doi: 10.1016/j.socscimed.2021.114195

31. Aizaki H, Fogarty J. An R package and tutorial for case 2 best–worst scaling. Journal of Choice Modelling. 2019;32.doi: 10.1016/j.jocm.2019.100171

32. Groemping U. DoE.base: Full Factorials, Orthogonal Arrays and Base Utilities for DoE Packages. [Internet]. 2017 [Available from:https://cran.r-project.org/web/packages/DoE.base/index.html].

33. Aizaki H. Support.BWS2: Basic Functions for Supporting an Implementation of Case 2 Best-Worst Scaling. [Internet]. 2019 [Available from:https://CRAN.R-project.org/package=support.BWS2.].

34. Therneau T. Survival: A Package for Survival Analysis in R [Internet]. 2021 [cited 2021 September 21]. Available from:https://CRAN.R-project.org/package=survival.].

35. Hosmer D, Hosmer T, Le Cessie S, et al. A Comparison of Goodness-of-fit Tests for the Logistic Regression Model. Statistics in Medicine. 1997;16:965–80.doi:

36. Hair J, Anderson R, Tatham R, et al. Multivariate Data Analysis. 3 ed. New York: Macmillan; 1995.

37. Peduzzi P, Concato J, Feinstein AR, et al. Importance of events per independent variable in proportional hazards regression analysis. II. Accuracy and precision of regression estimates. J Clin Epidemiol. 1995;48(12):1503–10.doi: 10.1016/0895-4356(95)00048-8

38. R Development Core Team. R: A Language and Environment for Statistical Computing. R Foundation for Statistical Computing Vienna, Austria2005 [Available from:http://www.R-project.org/].

39. Aiello AE, Coulborn RM, Perez V, et al. Effect of Hand Hygiene on Infectious Disease Risk in the Community Setting: A Meta-Analysis. American Journal of Public Health. 2008;98(8):1372–81.doi: 10.2105/ajph.2007.124610

40. Czeisler MÉ, Garcia-Williams AG, Molinari N-A, et al. Demographic Characteristics, Experiences, and Beliefs Associated with Hand Hygiene Among Adults During the COVID-19 Pandemic — United States, June 24–30, 2020. MMWR Morbidity and Mortality Weekly Report. 2020;69(41):1485–91.doi: 10.15585/mmwr.mm6941a3

41. Head KJ, Kasting ML, Sturm LA, et al. A National Survey Assessing SARS-CoV-2 Vaccination Intentions: Implications for Future Public Health Communication Efforts. Science Communication. 2020.doi: 10.1177/1075547020960463

42. Wadman M. Having SARS-CoV-2 once confers much greater immunity than a vaccine—but vaccination remains vital Science Insider2021 [Available from:https://www.science.org/content/article/having-sars-cov-2-once-confers-much-greater-immunity-vaccine-vaccination-remains-vital].

43. Atmar RL, Lyke KE, Deming ME, et al. Heterologous SARS-CoV-2 Booster Vaccinations - Preliminary Report. medRxiv. 2021.doi: 10.1101/2021.10.10.21264827

44. Wise J. Vaccinating against covid and flu at same time is safe, study shows. BMJ. 2021:n2411.doi: 10.1136/bmj.n2411

45. Leng A, Maitland E, Wang S, et al. Individual preferences for COVID-19 vaccination in China. Vaccine. 2021;39(2):247–54.doi: 10.1016/j.vaccine.2020.12.009

46. Ibuka Y, Li M, Vietri J, et al. Free-Riding Behavior in Vaccination Decisions: An Experimental Study. PLOS ONE. 2014;9(1):e87164.doi: 10.1371/journal.pone.0087164

47. Rieger M. Triggering altruism increases the willingness to get vaccinated against COVID-19. Social Health and Behavior. 2020;3(3).doi: 10.4103/shb.Shb_39_20

48. Cucciniello M, Pin P, Imre B, et al. Altruism and vaccination intentions: Evidence from behavioral experiments. Social Science & Medicine. 2021.doi: 10.1016/j.socscimed.2021.114195

49. Riva MA, Paladino ME, Paleari A, et al. Workplace COVID-19 vaccination, challenges and opportunities. Occupational Medicine (London). 2021.doi: 10.1093/occmed/kqab080

50. Magliozzi D, Saperstein A, Westbrook L. Scaling Up. Socius: Sociological Research for a Dynamic World. 2016;2.doi: 10.1177/2378023116664352

